# Disentangling Predictors of COPD Mortality with Probabilistic Graphical Models

**DOI:** 10.1101/2024.01.31.24301705

**Authors:** Tyler C. Lovelace, Min Hyung Ryu, Minxue Jia, Peter Castaldi, Frank C. Sciurba, Craig P. Hersh, Panayiotis V. Benos

## Abstract

**Background-Research question:** Chronic Obstructive Pulmonary Disease (COPD) is a leading cause of mortality. Predicting mortality risk in COPD patients can be important for disease management strategies. Although scores for all-cause mortality have been developed previously, there is limited research on factors that may directly affect COPD-specific mortality.

**Study design-Methods:** used probabilistic (causal) graphs to analyze clinical baseline COPDGene data, including demographics, spirometry, quantitative chest imaging, and symptom features, as well as gene expression data (from year-5).

**Results:** We identified factors linked to all-cause and COPD-specific mortality. Although many were similar, there were differences in certain comorbidities (all-cause mortality model only) and forced vital capacity (COPD-specific mortality model only). Using our results, we developed *VAPORED*, a 7-variable COPD-specific mortality risk score, which we validated using the ECLIPSE 3-yr mortality data. We showed that the new model is more accurate than the existing ADO, BODE, and updated BODE indices. Additionally, we identified biological signatures linked to all-cause mortality, including a plasma cell mediated component. Finally, we developed a web page to help clinicians calculate mortality risk using VAPORED, ADO, and BODE indices.

**Interpretation:** Given the importance of predicting COPD-specific and all-cause mortality risk in COPD patients, we showed that probabilistic graphs can identify the features most directly affecting them, and be used to build new, more accurate models of mortality risk. Novel biological features affecting mortality were also identified. This is an important step towards improving our identification of high-risk patients and potential biological mechanisms that drive COPD mortality.

## Introduction

Chronic Obstructive Pulmonary Disease (COPD) is a leading cause of mortality worldwide^1^. Predictive models of mortality in COPD can be used to identify high-risk individuals who may benefit from earlier or targeted interventions. As such, predictive models have been developed to predict all-cause mortality risk in individuals with COPD. Examples include the BODE (BMI, airflow obstruction, dyspnea, exercise capacity) index^2^ and its updated^3^ or expanded variants^4^, the ADO (age, dyspnea, airflow obstruction) index^3^, and the DOSE (dyspnea, airflow obstruction, smoking status, exacerbation frequency) index^5^. Of these, the ADO and updated BODE indices were found to perform best in a large-scale meta-analysis in external validation cohorts^6^. In addition to these simple clinical predictors, more complex machine-learning approaches that incorporate clinical, demographic, and imaging features have demonstrated improved prediction of all-cause mortality^7,8^.

These approaches have two key limitations which we seek to address. First, traditional machine learning methods, such as regression models^9,10^ and random survival forests (RFs)^11^, identify purely associative predictors and cannot provide insights into the possible causality of the observed interactions. In contrast, probabilistic graphs (also referred to as “causal graphs”)^12^ seek to learn potential cause-effect relationships in observational datasets^13^, by considering and factoring out any confounders. In biomedical settings, such approaches have been applied to identify direct effectors of an outcome and to develop efficient predictors^14,15^ or predict effects of interventions^16-18^. Here, we use a recently developed algorithm (**Supplementary Methods**) to construct probabilistic graph models from multi-modal data (*i.e.*, demographic, clinical, spirometry, chest CT scan, and biological features) to identify predictors that provide independent information for COPD-specific mortality.

Second, existing mortality predictors are trained, calibrated, and validated on all-cause rather than COPD-specific mortality of COPD patients. This introduces the possibility of incorrectly estimating the degree to which general risk factors contribute to COPD-specific mortality; or may introduce spurious associations due to the presence of comorbidities that act independently of the COPD-specific risk. Graph models, by construction, consider all confounder combinations and factor out the indirect effects and simple correlations. Here, we construct and compare separate graph models of all-cause and COPD-specific mortality. This allows us to disentangle features that are strongly independently informative to COPD-specific mortality from features informative of all-cause mortality. Furthermore, direct effectors identified by the graph models can be used to construct robust predictors of COPD-specific mortality risk. To further investigate blood-derived molecular signatures affecting mortality in COPD patients, we additionally construct graph models of all-cause mortality utilizing features derived from whole blood samples.

## Methods

### Study Population and Features

The discovery cohorts were derived from the COPD Genetic Epidemiology (COPDGene) Study, which recruited 10,198 current and former smokers, aged 45-80, from across US^19^. Various demographic, clinical, spirometry, and chest CT scan features were collected. Additionally, all-cause and COPD-specific mortality were recorded. A death was attributed to COPD if its cause was adjudicated to be COPD-related by the COPDGene criteria^20^ adapted from the TORCH UCD^21^. The features included in our mortality graph model analyses are key demographics (*e.g.*, age, sex, race), clinical measures (*e.g.*, BMI, resting oxygen saturation (SaO2)), measures of COPD-related physiology, and symptoms), and quantitative chest CT scan features (*e.g.*, %emphysema of the lungs^22^, segmental airway wall thickness (AWT)^22^). Relevant medical history and comorbidities (smoking status, pneumonia, diabetes) were also included. Our final **COPDGene Phase 1 study cohort** consisted of the 8,610 participants who had no missing values in the set of selected baseline features and longitudinal follow up data for all-cause and COPD-specific mortality.

COPDGene conducted a 5-yr follow-up study, during which, blood-derived biological features were measured (hemoglobin levels, platelet counts, white blood cell differential percentages, whole blood gene expression)^23^. To identify biological signatures linked to all-cause mortality in COPD, we constructed the **COPDGene Phase 2 study cohort**, that contains these blood-derived features in addition to the Phase 1 COPDGene measurements. The RNA-seq data was processed using CIBERSORTx^24^ and the leukocyte signature matrix (LM22)^25^ to infer more detailed cell type proportions. VIPER^26^ was used to infer transcription factor activity. This resulted in 3,182 participants with longitudinal follow up data for all-cause mortality, RNA-seq gene expression profiles, and no missing values in the set of selected features.

The ECLIPSE study^27^ was used for external validation of our findings. ECLIPSE has 2,312 COPD subjects or smoker controls that have 3-year mortality data, and no missing values among our selected features. **Table 1** presents the main characteristics of the cohorts, while **Suppl Table S1** presents the differences in characteristics between the two COPDGene cohorts.

**Table 1:**
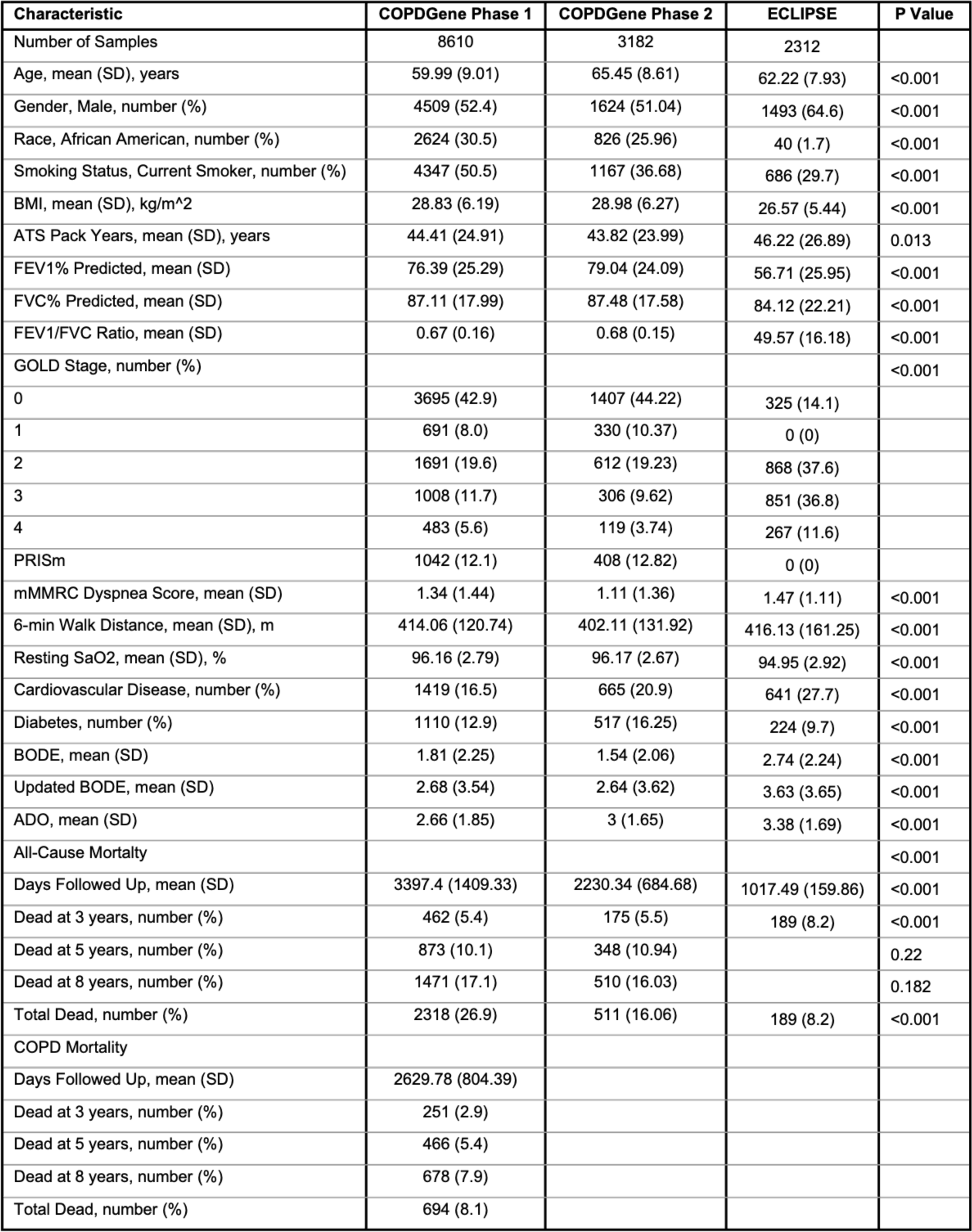
Clinical characteristics of the COPDGene Phase1 and Phase 2 studies used for the construction of graphical models of all-cause and COPD-specific mortality;, and the ECLIPSE 3-year all-cause mortality cohort, used for external validation.

### Construction of Graph-based Predictive Models

Directed probabilistic graph models (also referred to as “causal graph models”) for the COPDGene cohorts were constructed using the recently developed CausalCoxMGM method (**Supplementary Methods**). This method uses a two-step procedure to learn directed graphs from observational data in the presence of latent confounders. The resulting graphs can provide insights into potential mechanisms, generate hypotheses, and select minimal sets of the most relevant predictors (the direct neighbors only or the Markov blanket-MB)^28^. By construction, the MB variables of an outcome provide independent information to it.

We used both the direct neighbors and the full MB in the graphs to develop predictive Cox regression models^9^ of all-cause and COPD-specific mortality. These models were compared to the ADO and Updated BODE indices^3^, and to models learned with standard machine learning approaches: LASSO Cox Regression^29^ and RF^11^. Performance of these models was assessed through 5-fold cross-validation, with each model (except ADO and the updated BODE indices) trained on 80% of the data to predict the held-out 20% in each fold. Model performance was scored using the Harrell’s concordance index^30^, which represents the probability of a higher risk individual dying before a lower risk one. For models that perform feature selection (direct neighbors, MB, and LASSO Cox regression) we also compared the number of features each predictive model selected. For further details on causal graph discovery, model selection, and graph-based feature selection, see **Supplementary Methods**.

## Results

### Characteristics of Discovery and Validation Cohorts

As the Phase 2 study comprised from a subset of the Phase 1 study participants, there are some expected significant changes to clinical covariates (**Suppl Table S1**), such as an approximately five-year increase in age (p<0.001) and a statistically significant reduction of patients in more severe GOLD categories (p<0.001). We also observe significant increases in comorbidities’ incidence, such as cardiovascular disease (CVD; defined in ^8^) and diabetes (p<0.001). Among the standard mortality risk indices, there is a significant decrease in the BODE index^2^ and a significant increase in the ADO index^3^ of participants of Phase 2 compared to Phase 1 (p<0.001). Of particular interest is the distribution of all-cause mortality in the two phases. We see that there is not a statistically significant difference in the survival functions of the two phases, nor in the proportion of deaths at time points shared by both groups (3, 5, and 8 years). This is important as it suggests that, despite the older participants in Phase 2 and the exclusion of individuals who died before the five-year follow up, the all-cause mortality does not differ significantly between the two phases.

The ECLIPSE study^27^ differs significantly in its patient population compared to COPDGene (**Table 1**). It has significantly more male participants (p<0.001) and is less racially diverse (98.3% white). Additionally, ECLIPSE is enriched for more severe cases of COPD (GOLD Stages 2-4; p<0.001). This is also reflected in significant increases in the ADO, BODE, and updated BODE indices compared to COPDGene. Finally, likely due to the difference in COPD severity in the patient population, we see a significant difference in all-cause mortality between the two studies (p<0.001). This is also reflected in the number of deaths observed in the first three years, which is significantly higher in the ECLIPSE study (p<0.001).

### Comparison of All-Cause and COPD-specific Mortality Models Identifies Features Directly Linked to COPD Mortality

The MB variables (see **Supplementary Methods**) of all-cause mortality and COPD-specific mortality overlap substantially in COPDGene Phase 1. Overlapping features include classical predictors of mortality in COPD, such as age, 6MWD, mMRC Dyspnea score, BMI, and forced expiratory volume in 1s (FEV1)/forced vital capacity (FVC) ratio^3,31^ (**Figure 1, Supplementary Table S2**). Additionally, prior diagnosis of pneumonia, participant’s resting SaO2, AWT, history of severe exacerbations, and high blood pressure were linked to both all-cause and COPD-specific mortality. However, there were differences in the two models. FVC %predicted is strongly associated with COPD-specific mortality only, while FEV1 %predicted independently informs all-cause mortality only. Also, as expected, comorbidities such as CVD and diabetes were associated with all-cause mortality only; same applies to risk factors such as smoking status (at the visit), pack years, and gender. Finally, heart rate and the coughing up phlegm symptom are also linked to all-cause mortality only.

**Figure 1:**
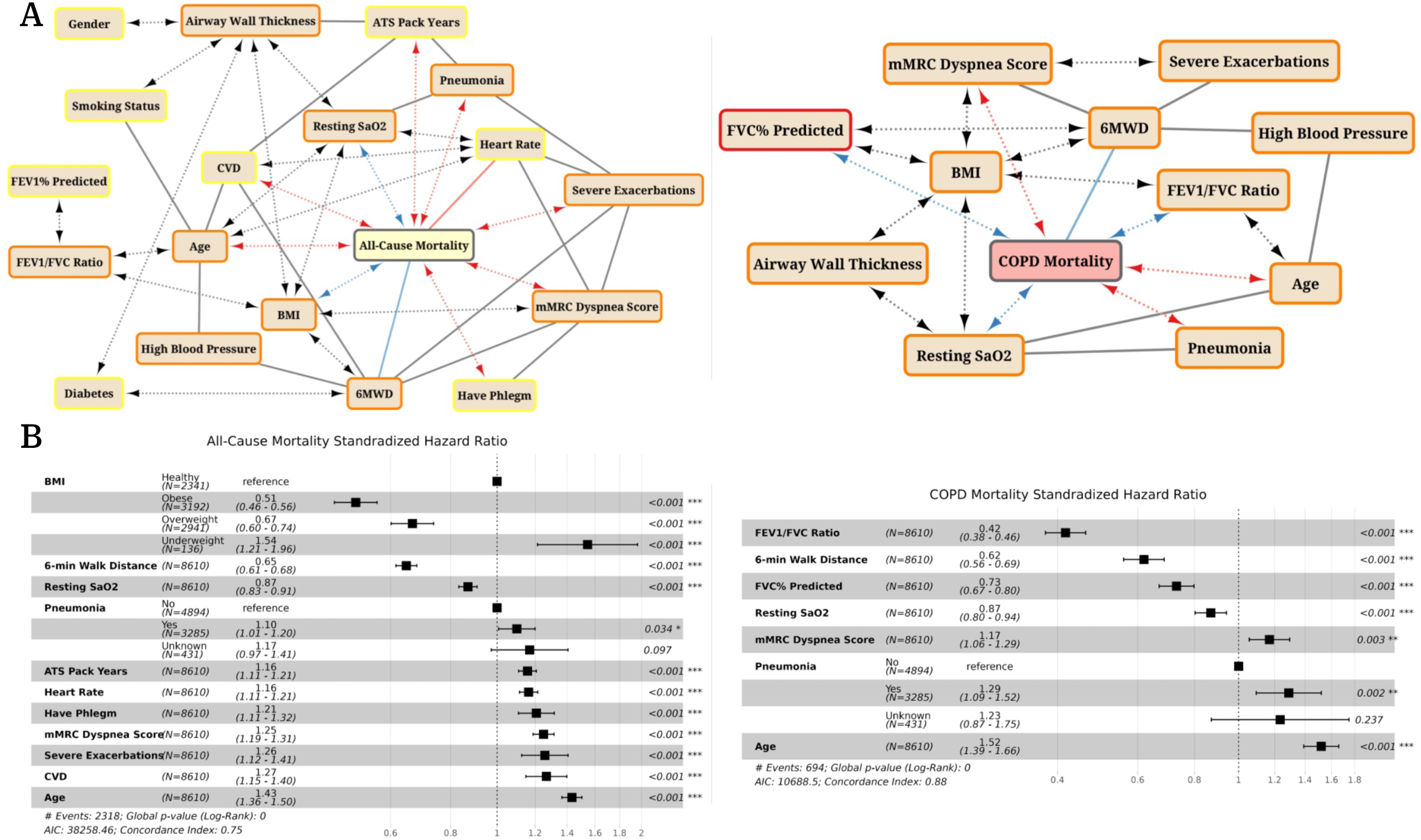
Graph models of all-cause (**A**, left) and COPD-specific (**A**, right) mortality in the COPDGene Phase 1 study. Only the Markov blanket (MB) mortality features are shown. Node boundaries are colored to represent whether a feature is present in the MB of all-cause mortality (yellow), COPD-specific mortality (red), or both (orange). Adjacencies in the graphical model represent a direct interaction between two variables, while edge orientations represent the type of interaction inferred by CausalCoxMGM. Undirected edges (X --- Y) represent a direct interaction between X and Y, while bidirected edges (X <-> Y) mean that some unobserved confounder affects X and Y. Edge color designates whether higher values of this MB variable represent lower (blue) or (red) higher mortality risk. The standardized hazard ratios of the direct neighbors all-cause (**B**, left) and COPD-specific mortality (**B**, right) display the direction, relative effect size, and significance of each covariates’ association with mortality. Hazard ratios for numeric features represent the change in hazard for a 1 SD increase. Although all depicted MB variables contribute independent information to each mortality variable, we observed no significant decrease in prediction accuracy in models with only the direct neighbors.

Looking only at the direct neighbors of COPD-specific mortality provides some additional insights. BMI, although it provides some independent information for COPD-specific mortality, is not directly linked to it, suggesting that its association with mortality in COPD is through its interactions with FEV1/FVC, FVC %predicted, 6MWD, mMRC Dyspnea score, and resting SaO2 (**Figure 1A**, right). However, BMI is directly linked to all-cause mortality. Additionally, we observe a direct effect of FVC %predicted to COPD-specific but not to all-cause mortality. **Figure 1B** shows the relative importance (hazard ratios) of each direct neighbor to mortality in the two graphs, which are all independently significant.

### Predictive Models of COPD Mortality

In addition to providing insights on direct interactions in observational datasets, graph models are useful tools for constructing robust, parsimonious, and powerful predictive models. We found that the graph-based predictive models (Neighbors, MB) significantly outperformed the ADO and updated BODE indices for both all-cause and COPD-specific mortality (**Figure 2A**). The models based on direct neighbors only (i.e., excluding “spouses” from the MB) performed similarly to models constructed from the full MB, LASSO Cox regression, and RF, but consistently selected significantly fewer features. The predictive models based on the full MB performed similarly to LASSO Cox regression and RF, while still being significantly more parsimonious than LASSO.

**Figure 2:**
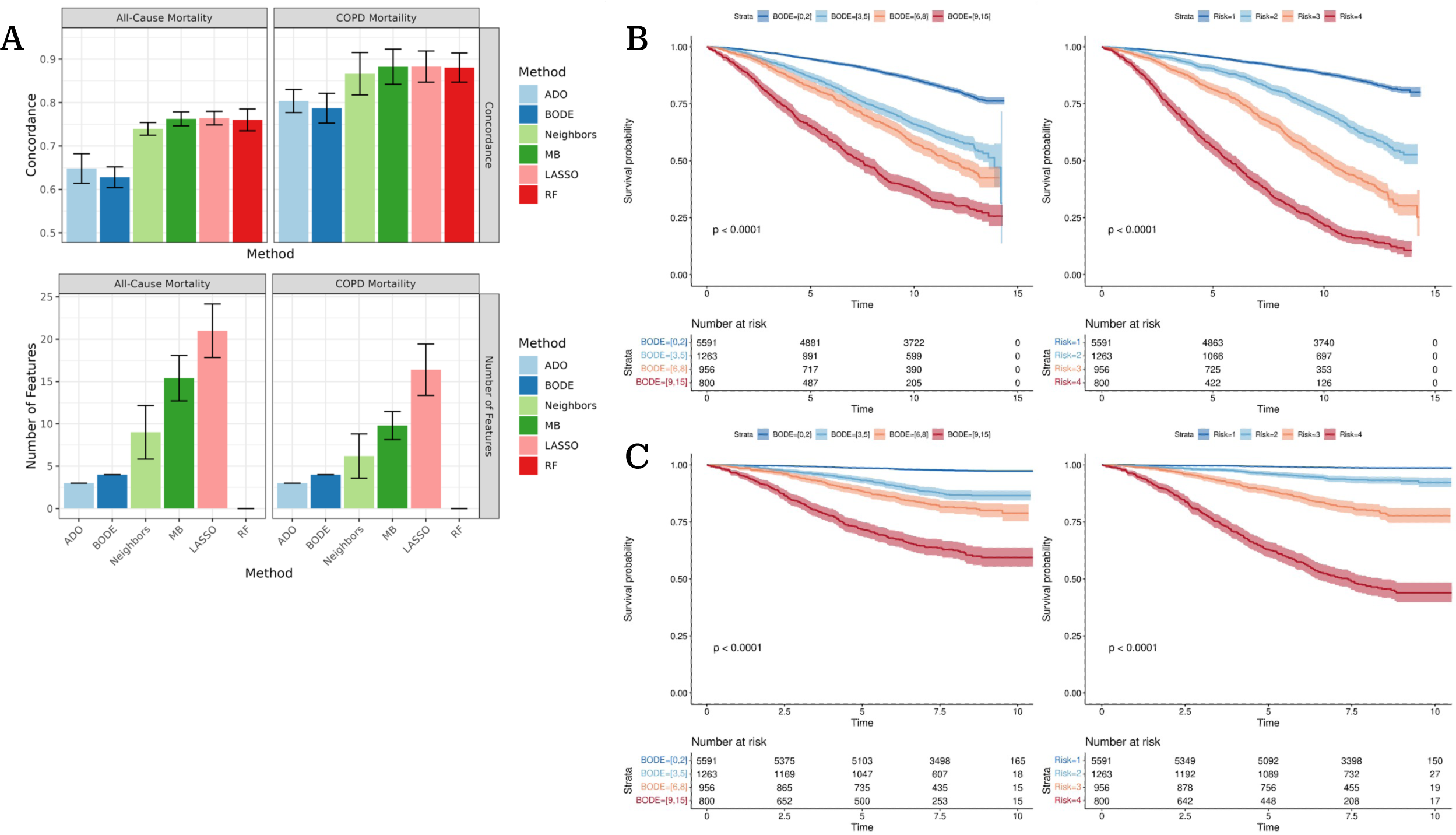
Performance of predictive models of all-cause and COPD-specific models assessed through 5-fold cross-validation (**A**). Models constructed using the direct neighbors of mortality (Neighbors) and all Markov blanket variables (full MB) were compared against the ADO and updated BODE indices, as well as LASSO Cox regression and random survival forests (RF) models trained on the same cross-validation splits. Model performance was assessed via Harrell’s concordance. The number of features for models where feature selection was performed (Neighbors, full MB, LASSO Cox) was also assessed. Error bars denote the 95% confidence intervals of the cross-validation estimates. To demonstrate the ability of the full MB models to stratify patients by risk of all-cause (**B**) and COPD-specific (**C**) mortality compared to the updated BODE index, we stratify individuals into four risk groups using the updated BODE index (left) as well as four risk groups of equal size predicted by the full MB models (right). Confidence bands represent the 95% confidence interval.

To demonstrate the ability of our models to stratify patients better than the updated BODE index, we created four risk groups based on the updated BODE score as well as groups of equal size from our full MB model. The Kaplan-Meier survival probability estimates^32^ were plotted for these four risk groups for both all-cause (**Figure 2B**) and COPD-specific mortality (**Figure 2C**). The new full MB risk groups stratify patients into groups with significantly different survival curves for both all-cause and COPD-specific mortality. Additionally, COPD-specific mortality was considerably easier to predict compared to all-cause mortality (**Figure 2BC**).

### External Validation: the ECLIPSE Study

In our model, COPD-specific mortality has seven direct neighbors (**Supplementary Table S2**, bold), which are easily obtainable clinical measurements (no need for chest CT scans), and they are frequently measured across COPD studies. We trained a predictor model of COPD-specific mortality with these seven variables from COPDGene Phase 1 and constructed an index of COPD mortality risk as in ^3^ with the categories presented in **Table 2**. Our model, *VAPORED* (**<u>V</u>**ital capacity-FVC %predicted, **<u>A</u>**ge, history of **<u>P</u>**neumonia, **<u>O</u>**xygen saturation, the FEV1/FVC **<u>R</u>**atio, 6-min walk <u>**E**</u>xercise capacity, **<u>D</u>**yspnea), was validated on the 3-year mortality data in the ECLIPSE study and compared to the standard ADO, BODE, and updated BODE indices. We used four measures to assess predictive power: concordance, the concordance probability estimate (CPE), the cumulative/dynamic (C/D) AUC at 3 years, and the integrated C/D AUC. Means and standard errors of these metrics were estimated over 2000 bootstrapped samples. While the ADO, BODE, and Updated BODE indices have similar predictive power across all metrics, *VAPORED* had consistently the highest predictive power by at least one standard error (**Figure 3A**) and significantly higher CPE than all other indices (p<0.05). All-cause mortality survival functions for each *VAPORED* score interval were estimated by training a parametric Cox regression model with a Weibull baseline hazard on COPDGene Phase 1 data^10^ (**Supplementary Figure S1A**; **Supplementary Table S3**). In **Supplementary Figure S1B** we see that the predictions of this model are not significantly different than the observed survival probabilities for each calibration plot in 1-year, 2-years and 3-years survival. Patients in the ECLIPSE dataset were also stratified into four risk groups according to the BODE score and four risk groups of equivalent size based on the *VAPORED* score (**Figure 3B**). The *VAPORED* score categories have significantly different survival probability in the ECLIPSE study and *VAPORED* shows greater separation amongst risk groups than the BODE score.

**Figure 3:**
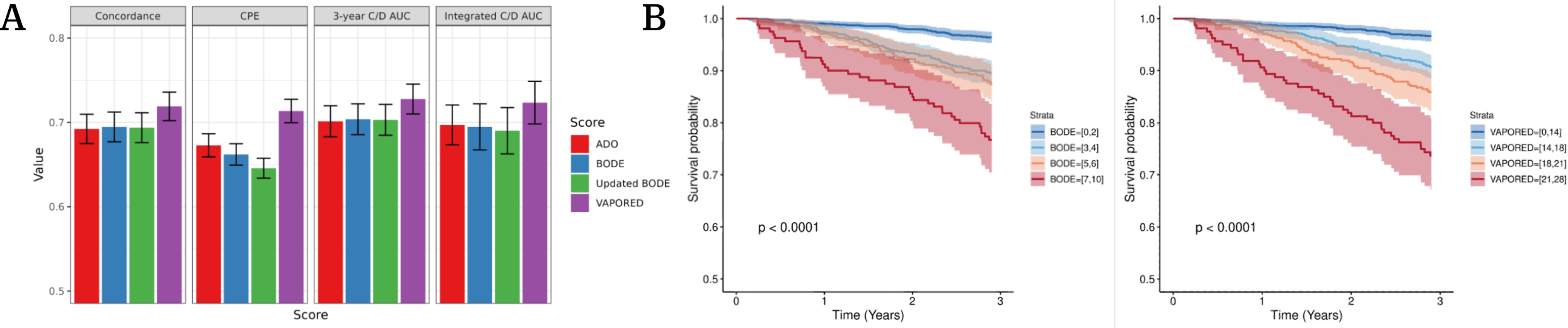
Performance of the VAPORED risk score in the ECLIPSE 3-year all-cause mortality external validation cohort. The concordance, concordance probability estimate (CPE), 3-year C/D AUC, and integrated C/D AUC for the VAPORED, ADO, BODE, and updated BODE scores (**A**). Error bars denote the 95% confidence intervals of the bootstrapped estimates. To demonstrate the ability of the VAPORED risk score to stratify patients by risk of all-cause mortality in the ECLIPSE study compared to the updated BODE index (**B**), we stratify individuals into four risk groups using the BODE index (left) as well as four risk groups of approximately equal size by VAPORED risk score (right). Confidence bands represent the 95% confidence interval.

**Table 2:** Scoring criteria for VAPORED mortality score, constructed from the direct neighbors of COPD-specific mortality in the graphical model.

| VAPORED Risk Score |  |  |
| --- | --- | --- |
| Feature | Category | Score |
| Age (years) | < 50 | 0 |
|  | [50, 60) | 1 |
|  | [60, 70) | 3 |
|  | [70, 80) | 5 |
|  | ≥ 80 | 6 |
| 6-min Walk Distance (m) | ≥ 550 | 0 |
|  | [350, 550) | 2 |
|  | [250, 350) | 4 |
|  | [150, 250) | 5 |
|  | < 150 | 7 |
| FEV1/FVC Ratio (%) | ≥ 80 | 0 |
|  | [65, 80) | 3 |
|  | [50, 65) | 5 |
|  | [35, 50) | 6 |
|  | < 35 | 8 |
| FVC% Predicted (%) | ≥ 100 | 0 |
|  | [80, 100) | 1 |
|  | [60, 80) | 3 |
|  | [45, 60) | 4 |
|  | < 45 | 5 |
| mMRC Dyspnea score | [0, 1] | 0 |
|  | 2 | 1 |
|  | [3, 4] | 2 |
| Pneumonia | No / Unknown | 0 |
|  | Yes | 1 |
| Resting SaO2 (%) | ≥ 93 | 0 |
|  | [85, 93) | 2 |
|  | < 85 | 3 |

### Graph-based Model of All-Cause Mortality in COPD Reveals Biological Signatures of Mortality Risk

In addition to the demographic, clinical, spirometry, and chest CT scan features, COPDGene Phase 2 Study also collected biological data, such as white blood cell differential percentages, hemoglobin levels, and whole blood gene expression. This provides a unique opportunity to investigate how informative the biological features are in the context of the clinical features and comorbidities. We applied our graph-modeling approach to identify features potentially affecting all-cause mortality in this dataset. We found eight of the 18 previously identified clinical features in the MB (age, gender, BMI, 6MWD, mMRC Dyspnea score, FEV1/FVC, heart rate, CVD; **Supplementary Table S2**). Additional variables linked to all-cause mortality include: a socioeconomic feature, *Internet Access*, which was not collected during Phase 1, and five biological features (platelets, hemoglobin, SPIB transcription factor activity, and CIBERSORTx inferred proportions of plasma cells and resting memory CD4+ T cells) (**Figure 4AB**). We used 5-fold cross-validation (internally) to ensure that our graph modeling approach was learning robust sets of features of all-cause mortality. As above, the model based on the identified predictors of all-cause mortality significantly outperformed the ADO and updated BODE indices in terms of concordance, while it is significantly more parsimonious than the other machine learning methods (LASSO Cox regression, RF) without a significant loss in predictive accuracy (**Figure 4C**). Finally, the COPDGene Phase 2 full MB predictive model can stratify individuals into significantly different mortality risk groups (**Figure 4D**). These results strengthen the claim that our method derives relevant direct interactions between features from this multi-modal and multi-scale dataset and all-cause mortality.

**Figure 4:**
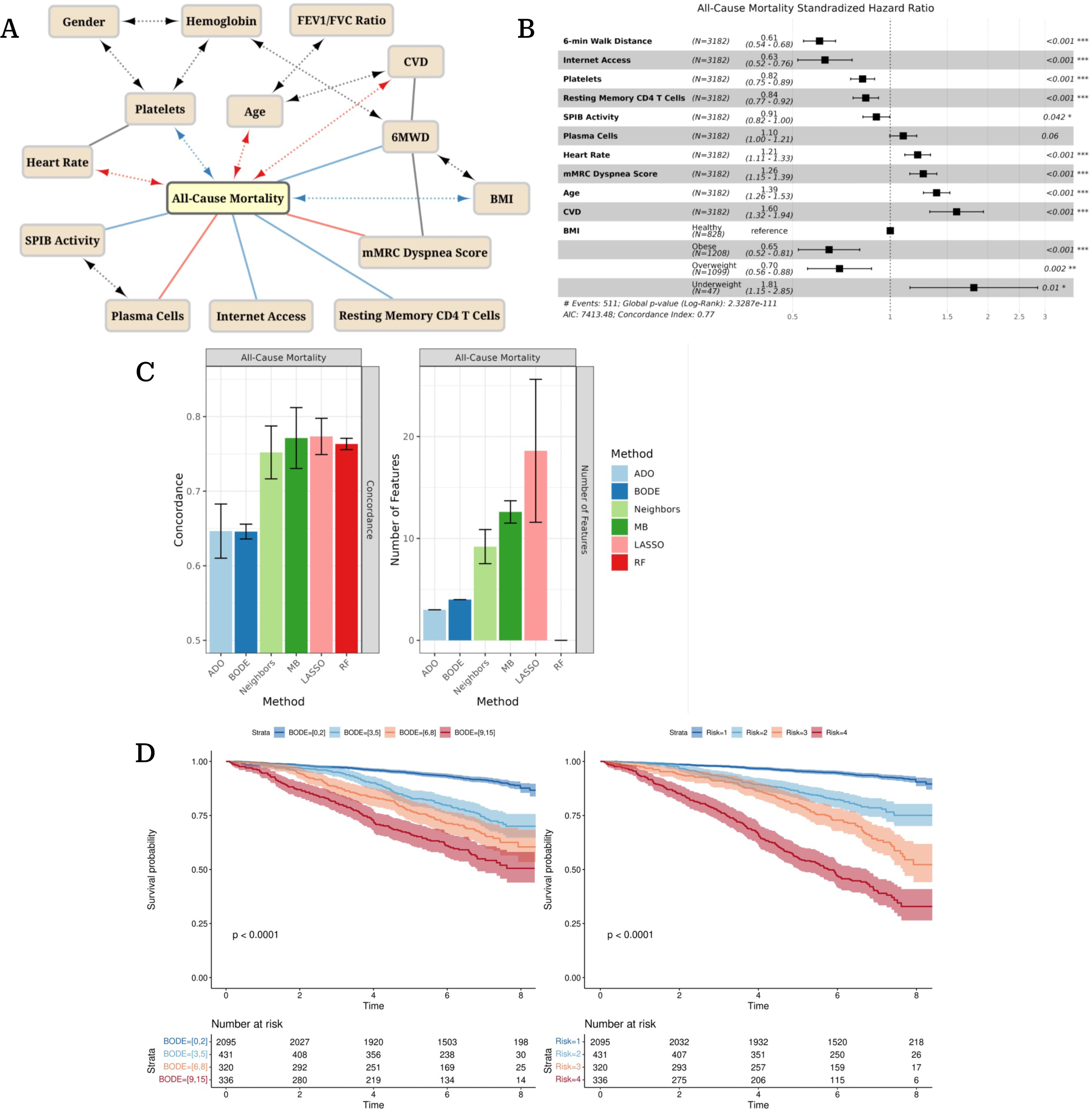
Graph models of all-cause mortality in the COPDGene Phase 2 study (**A**). Only the Markov blanket (MB) of all-cause mortality is shown. Adjacencies in the graphical model have the same notation as in Figure 1. The standardized hazard ratios of the direct neighbors all-cause mortality (**B**) display the direction, relative effect size, and significance of each covariates’ association with mortality. Hazard ratios for numeric features represent the change in hazard for a 1 SD increase. The performance of predictive models of all-cause and COPD-specific models assessed through 5-fold cross-validation (**C**). Models constructed using the direct neighbors of mortality (Neighbors) and Markov blanket of mortality (full MB) were compared against the ADO and updated BODE indices, as well as LASSO Cox regression and random survival forests (RF) models trained on the same cross-validation splits. Model performance was assessed via Harrell’s concordance and the number of features for models where feature selection was performed (Neighbors, full MB, LASSO Cox). Error bars denote the 95% confidence intervals of the cross-validation estimates. To demonstrate the ability of the full MB model to stratify patients by risk of all-cause mortality compared to the updated BODE index (**D**), we stratify individuals into four risk groups using the updated BODE index (left) as well as four risk groups of equal size using full MB model predictions (right). Confidence bands represent the 95% confidence interval.

### Web-based tool for all-cause and COPD-specific mortality

To help people further evaluate our VAPORED score predictor, we developed a web-based tool. The web tool allows the user to input the seven VAPORED key variables (FVC %predicted, age, history of pneumonia, SaO2, FEV1/FVC ratio, 6MWD, mMRC Dyspnea score) for an individual and outputs two mortality risk curves (all-cause, COPD-specific) for the next 10 years. In addition, if the user provides values for BMI and FEV1 %predicted, the web tool outputs similar curves for BODE and ADO risk scores (for comparison purposes). The tool is available as a Shiny app from: https://vapored.shinyapps.io/VAPORED/. Some example values have been pre-loaded.

## Discussion

In this study, we applied a probabilistic graph modeling approach to distinguish features directly linked to COPD-specific and all-cause mortality from simple correlates. We analyzed demographic, clinical, spirometry, and chest CT scan features from baseline COPDGene measurements. The graphical models, by construction, are considering all possible combinations of covariates and filter out the indirect effects.

### Key results – Interpretation

We found known risk factors of all-cause mortality in COPD to inform both models. These include age, mMRC Dyspnea score, 6MWD, and BMI, which are used in BODE and ADO indices. Interestingly, these features remained informative of all-cause mortality even after biological data were added to the model (from COPDGene Phase 2). Other common features of the all-cause and COPD-specific mortality baseline models (history of pneumonia, and resting SaO2) did not appear in the Phase 2 model, probably because their information is superseded by the biological variables of this model (Platelets, Resting Memory CD4+ T Cells, SPIB Activity, and Plasma Cells). Further investigation is needed to determine potential long-term biological effects of pneumonia to COPD patients.

The all-cause and COPD-specific mortality models have some unique characteristics, despite their substantial overlap. FVC %predicted appeared only in the COPD-specific model. This might indicate the effect of hyperinflation (i.e., residual volume) in COPD mortality, which was recently shown to be better represented by FVC %predicted and FEV1/FVC rather than FEV1 %predicted^33^. Further, hyperinflation is more strongly linked to mortality than FEV1^34,35^. FEV1/FVC ratio, which is independent on race specific reference equations, is a better discriminator of mortality than FEV1 %predicted^36^. Not surprisingly, certain comorbidities are informative for the all-cause mortality only: diabetes, CVD, heart rate. Pack years, which affects multiple systems, is informative for the all-cause mortality, but not the COPD-specific mortality. This is probably because direct measurement of impacted lung variables has incorporated the smoking information. Finally, the important contribution of pneumonia to our COPD specific model is unique, although not unexpected given the established relationship between pneumonia and mortality in patients with COPD and the potential that pneumonia may cause impairment in lung immunity not reflected in the other metrics^37,38^.

The graph models enabled us to develop a new, parsimonious but informative, 7-feature risk score for COPD-specific mortality consisting of easily obtainable characteristics (age, 6MWD, FEV1/FVC, FVC %predicted, mMRC Dyspnea score, history of pneumonia, resting SaO2). We validated this new model in the ECLIPSE 3-year all-cause mortality data, as ECLIPSE has not recorded COPD-specific mortality. Our score consistently outperformed the ADO, BODE, and updated BODE indices across multiple metrics and had a significantly higher mortality CPE.

We also took advantage of COPDGene 5-year follow-up data, which additionally included socioeconomic factors and biological measurements. Our graph approach identified potentially important clinical and biological factors of all-cause mortality in current and former smokers with or at high risk of developing COPD. Age, BMI, FEV1/FVC, mMRC, 6MWD were still connected to all-cause mortality, as well as comorbidities CVD and heart rate. In addition, *Internet Access* was directly linked to all-cause mortality. Further investigation is needed to determine whether this is a surrogate for income or rural *vs* urban population characteristics.

Five biological features were linked to all-cause mortality in the Phase 2 model: hemoglobin levels, platelet counts, SPIB transcription factor activity, plasma cell proportions, and resting memory CD4+ T cell proportions. Low hemoglobin levels were associated with an increased risk of all-cause mortality, as has been previously observed in a population-based study of individuals with COPD^39^ and in patients who were admitted to hospital for COPD^40^. Platelets have also been previously associated with all-cause mortality in COPD, and antiplatelet therapies have been shown to reduce all-cause mortality in individuals with COPD^41,42^. This contradicts the hazard ratio learned by our model, which suggests that low platelet levels are associated with increased all-cause mortality. However, previous analysis has found a U-shaped association of platelet counts with all-cause mortality in COPD^43^. This indicates that both high and low platelet counts are associated with a higher risk of all-cause mortality, and the hazard ratio observed in our model is likely due to limitations of our modeling assumptions, which expect such associations to be monotonic. Even so, our graph discovery algorithm has correctly identified a biological signature that has been previously identified as having a likely causal effect on all-cause mortality in COPD patients^41,42^.

Regarding mechanistic insights, our model suggests that the plasma cell proportion and SPIB transcription factor activity are affecting mortality. The two likely interact (our model supports that), as the SPIB transcription factor is a negative regulator of plasma cell differentiation and immunoglobulin production^44,45^. Previous studies have linked B cell activity and the humoral immune response with COPD progression^46^. The formation of lymphoid follicles in the lung^46,47^ and larger numbers of infiltrating B cells, memory B cells, and plasma cells are associated with COPD severity^46^. Our model provides support for this mechanism of COPD progression, as elevated plasma cells and reduced SPIB transcription factor activity are found to be associated with an increased risk of all-cause mortality.

Our graph model also suggests that lower resting memory CD4+ T cells directly affect increase in all-cause mortality, independently of age or sex. Interestingly, circulating memory CD4+ T cells are known to increase with age^48^, and smoking^49,50^, and have been linked to an increase in IL-22 secretion, which has been previously implicated in COPD pathogenesis^51^. However, independently of these known risk factors for all-cause mortality in COPD, our results suggest that memory CD4+ T cells play a protective role in patients with COPD. A previous study in patients with COPD found that CD4+ T cell cytokine production in response to stimulus is restricted almost entirely to memory CD4+ T cells^52^. In healthy populations, memory CD4+ T cell populations can respond faster and more efficiently to previously experienced infections^48,53^. In both healthy individuals and those with COPD, circulating and tissue resident memory CD4+ T cells that respond to common viral respiratory pathogens were found, without any significant defects between the COPD and control subjects^54^. The association of low levels of resting memory CD4+ T cells with an elevated risk of all-cause mortality observed in our analysis may reflect the importance of memory CD4+ T cell populations in the response to respiratory infections in subjects with COPD.

### Limitations

The observational nature of both cohorts makes difficult to establish true causal relationships. Additionally, the COPDGene Study contains older individuals with extensive smoking histories, so the *VAPORED* score reflects this. Thus, its application should be limited to individuals who have or are at high risk of developing COPD. Finally, the modeling assumptions of our probabilistic graph models (additive monotonic relationships, Markov faithfulness) place limitations on the interactions our models can recover. Despite these limitations, our approach had similar predictive power compared to standard indices or common machine learning methods for mortality prediction.

## Conclusions

Our graph-based models can go beyond simple correlates and identify direct effectors of outcomes. It is also important to distinguish COPD-specific from all-cause mortality, a subject that has been understudied in the past. We developed a new COPD-specific mortality risk score (*VAPORED*), which is significantly better than established risk scores, and we validated our findings in an external cohort. Furthermore, we identified socioeconomic and biological factors that contribute to all-cause mortality in COPD patients. In the future, we plan to extend this study to additional modalities, such as genetic information, blood proteomics or methylomics, and develop truly comprehensive mortality risk scores. Accurate risk stratification of COPD patients can aid in the identification of high-risk individuals who may benefit most from targeted interventions.

## Data Availability

No new data were produced during this study. The COPDGene and ECLIPSE data can be obtained from dbGAP.
- COPDGene: https://www.ncbi.nlm.nih.gov/projects/gap/cgi-bin/study.cgi?study_id=phs000179.v1.p1
- ECLIPSE: https://www.ncbi.nlm.nih.gov/projects/gap/cgi-bin/study.cgi?study_id=phs001472.v2.p1

https://www.ncbi.nlm.nih.gov/projects/gap/cgi-bin/study.cgi?study_id=phs001472.v2.p1

## Acknowledgement

This work was supported by NHLBI grants R01HL159805 and R01HL157879 (to PVB) and F31LM013966 (to TCL). This work was also supported by NHLBI U01HL089897 and U01HL089856. The COPDGene study (NCT00608764) is also supported by the COPD Foundation through contributions made to an Industry Advisory Committee comprised of AstraZeneca, Bayer Pharmaceuticals, Boehringer-Ingelheim, Genentech, GlaxoSmithKline, Novartis, Pfizer and Sunovion.

## Abbreviations

MB: Markov blanket
RF: Random Forest

